# Key factors underpinning neuroimmune-metabolic-oxidative (NIMETOX) major depression in outpatients: paraoxonase 1 activity, reverse cholesterol transport, increased atherogenicity, protein oxidation, and differently expressed cytokine networks

**DOI:** 10.1101/2025.03.02.25323183

**Authors:** Michael Maes, Ketsupar Jirakran, Laura de Oliveira Semeão, Ana Paula Michelin, Andressa K. Matsumoto, Francis F. Brinholi, Decio S. Barbosa, Chavit Tivirachaisakul, Abbas F. Almulla, Drozdstoj Stoyanov, Yingqian Zhang

## Abstract

**Background:** Major depressive disorder (MDD) is associated with neuro-immune – metabolic – oxidative (NIMETOX) pathways.

**Aims:** To examine the connections among NIMETOX pathways in outpatient MDD (OMDD) with and without metabolic syndrome (MetS); and to determine the prevalence of NIMETOX aberrations in a cohort of OMDD patients.

**Methods:** We included 67 healthy controls and 66 OMDD patients and we assessed various NIMETOX pathways.

**Results:** We successfully identified a subgroup of individuals with aberrations in NIMETOX pathways, including diminished lecithin-cholesterol acyltransferase (LCAT), paraoxonase 1 (PON1) activity, and reverse cholesterol transport (RCT) activities, and elevated atherogenicity, differentially expressed immune networks, and advanced oxidation protein products (AOPP). A large part of the variance (around 44%) in atherogenicity indices was associated with AOPP, fasting blood glucose (FBG), PON1 activity, and immune activation. LCAT activity was positively correlated with PON1 activity and negatively with FBG, AOPP and immune activation. RCT was positively related with the PON1 R/R 192 genotype and negatively with FBG and immune activation. A larger part of the variance in the overall severity of OMDD (50.4%), suicidal behaviors (27.7%), and neuroticism (42.1%) was positively associated with adverse childhood experiences and NIMETOX pathways, including AOPP, immune-related neurotoxicity, FBG, insulin, and atherogenicity, and inversely with immune-related neuroprotection.

**Conclusions:** Many OMDD patients (78.8%) show aberrations in NIMETOX pathways. The features of OMDD, including severity of illness, neuroticism, and suicidal behaviors, are caused by intertwined NIMETOX pathways that may exert additional effects depending on whether MetS is present or not.

## Introduction

Major depressive disorder (MDD) is linked to intertwined anomalies in the neuro-immune system, metabolic disorders, and oxidative stress, collectively referred to as the NIMETOX pathways (including <u>n</u>euro-immune, <u>met</u>abolic, and <u>ox</u>idative factors) [1–8]. The acute phase of severe MDD is marked by the activation of the immune-inflammatory response system (IRS) and the compensatory immunoregulatory system (CIRS), which mitigates the IRS to avert hyperinflammation [9]. IRS activation is more significant than CIRS activation during the acute phase of severe inpatient MDD [10]. Central components in this IRS response are interleukin-16 (IL-16) and tumor necrosis factor (TNF)-signaling, as seen by elevated levels of TNF-α, TNF-β, and TRAIL (TNF-related apoptosis-inducing ligand [10].

The acute episode of MDD consists of two qualitatively separate categories: Major Dysmood Disorder (MDMD) and Simple Dysmood Disorder (SDMD) [11, 12]. MDMD is defined by a) heightened recurrence of illness (ROI), encompassing the frequency of depressive episodes and suicidal behaviors, elevated neuroticism scores, and a greater prevalence of dysthymia and anxiety disorders (collectively referred to as DYSNEUANX; and b) augmented severity of the current phenotype, conceptualized as a latent trait derived from the severity of depression, anxiety, and suicidal behaviors [11, 12]. Crucially, acute MDMD is marked by IRS activation in contrast to CIRS activation, whereas SDMD is associated with a suppression of CIRS, as seen by diminished activity of key immunoregulatory cytokines, including IL-4, IL-10, soluble sIL-2 receptor (sIL-2R), and IL-12p40 [12, 13].

Outpatient MDD (OMDD) exhibits a unique immune profile compared to MDMD and SDMD [14]. Firstly, OMDD is characterized by a closely interconnected immune-related neurotoxic (IM-NT) network featuring heightened levels of TNF-β and TNF signaling, IL-4, IL-9, and sIL-1RA, along with the chemokines CXCL12, CCL2, CCL4, and CCL11, as well as platelet-derived growth factor (PDGF). This indicates an augmented neurotoxic potential attributable to oxidative stress, neurodegeneration, cytotoxicity, cell death processes, disruption of the blood-brain barrier (BBB), and diminished neurogenesis [14]. Secondly, OMDD is characterized by the downregulation of vascular endothelial growth factor (VEGF), IL-12, CCL3, macrophage colony-stimulating factor (M-CSF), IL-1β, and nerve growth factor (NGF), signifying diminished neurogenesis and regulation of neuronal death [14].

Additional biomarkers of MDMD, SDSMD, and/or OMDD include elevated oxidative and nitrosative stress, diminished indicators of lecithin-cholesterol acyltransferase (LCAT) activity and reverse cholesterol transfer (RCT) activity, and heightened atherogenicity [5–8, 15]. Therefore, these three subtypes of depression exhibit a diminished RCT, a critical detoxification pathway that eliminates free cholesterol from the body, thus averting atherogenicity and safeguarding against heightened oxidative processes and IRS responses [16–19].

The primary factors contributing to the diminished activity of RCT in MDD are decreased levels of high-density lipoprotein (HDL) cholesterol, reduced LCAT activity, and lower apolipoprotein A (ApoA) levels [2, 5, 6, 7, 8]. LCAT facilitates RCT by transforming free cholesterol into cholesteryl esters on HDL particles, and its activity can be assessed by evaluating the esterification rate using free cholesterol assays [5, 7]. In OMDD, data suggests that reduced RCT correlates with heightened atherogenicity, as seen by an elevated atherogenicity index of plasma (triglycerides/HDL), Castelli risk index 1 (total cholesterol/HDL), and the ApoB/ApoA ratio [14]. Elevated ApoE levels contribute to heightened atherogenicity in OMDD and exacerbate the severity of depressive phenotypes, including suicidal behaviors [14].

Similarly, reduced RCT in MDD/OMDD may partially stimulate IM-NT network activation [8, 14] and, given its significance as a primary antioxidant mechanism, elevate oxidative stress [20, 21]. MDD is marked by elevated lipid and protein peroxidation, aldehyde production, and diminished lipid-associated antioxidant defenses [5, 15, 20, 21]. MDD is associated with elevated generation of nitric oxide metabolites (NOx), potentially leading to heightened nitrosylation observed in MDD [20, 21]. Furthermore, paraoxonase 1 (PON1), an antioxidant enzyme with antiatherogenic and anti-inflammatory properties [5, 22–24] is significantly decreased in MDD and associated with the features of MDD including increasing ROI [5, 22, 23, 25]. However, in OMDD, there is no data indicating whether elevated protein oxidation, as seen by plasma levels of advanced oxidation protein products (AOPPs), and lowered PON1 enzyme activities or alterations in PON1 Q192R gene variants correlate with atherogenicity indices and the clinical characteristics of OMDD.

MDD and metabolic syndrome (MetS) demonstrate a notable comorbidity, with their concurrent occurrence linked to increased atherogenicity and insulin resistance in association with oxidative and nitrosative stress (review: [3, 4, 26–28]. Elevated levels of inflammatory markers indicate that MetS may be linked to a mild chronic inflammatory response [29–33]. It was previously shown that in OMDD, the presence of MetS enhances TNF-signaling, IM-NT potential, and reduces immune-related neuroprotection (IM-NP) [32]. In OMDD, there are significant correlations between oxidative stress biomarkers, including AOPP, on the one hand, and atherogenicity and insulin resistance (based on fasting blood glucose or FBG and insulin assessments), on the other [28].

Significantly, we noted that the correlations between immunological and atherogenicity biomarkers and the clinical characteristics of OMDD varied considerably across patients with and without MetS [14, 32]. Consequently, whereas no significant correlations between RCT or atherogenicity markers and clinical OMDD characteristics were observed in participants with MetS, robust connections were identified in subjects without MetS [14]. The IM-NT indicators exhibited a robust correlation with clinical characteristics in the MetS group, but not in the non-MetS group [32]. However, no studies have investigated the interactions of NIMETOX pathways in individuals with OMDD and how these interactions are associated with the clinical characteristics of the condition. Moreover, no studies have investigated the potential identification of a subpopulation of patients with activated NIMETOX pathways and OMDD among outpatient populations with depression. Hence, this study aimed to examine (1) the connections among NIMETOX factors in OMDD and persons with and without MetS; and (2) whether a cohort of patients exhibiting elevated NIMETOX biomarkers and clinical characteristics of OMDD may be identified.

## Methods and Participants

### Participants

All subjects willingly submitted written informed consent before their participation in the study. The research was executed in accordance with international ethical standards and Thai privacy legislation. The Institutional Review Board of the Faculty of Medicine at Chulalongkorn University in Bangkok, Thailand, approved the inquiry under authorization number #445/63. This study included sixty-seven healthy controls and sixty-six individuals with OMDD. The control group was recruited through oral communication within the catchment area of Bangkok, Thailand. Patients with OMDD were recruited as outpatients from the Department of Psychiatry at King Chulalongkorn Memorial Hospital in Bangkok, Thailand. To assess the influence of MetS on the relationships between biomarkers and clinical data, we selected approximately 50% of the control and OMDD samples to be diagnosed with MetS. Consequently, sixty-four subjects were classified with MetS, whereas sixty-nine did not have the condition. Out of sixty-seven controls, thirty-three were diagnosed with MetS, while among sixty-six OMDD patients, thirty-one were diagnosed with MetS.

The outpatients were diagnosed with MDD according to the criteria specified in the DSM-5. Our OMDD sample accurately reflects the Bangkok OMDD population for the following reasons: a wide recurrence of illness (ROI); index spanning from 1 to 20 episodes; a comprehensive Hamilton Depression Rating Scale (HAMD) score range of 7 to 33 (encompassing non-responders and partial remitters to treatment); a female to male sex ratio of 2.66; and a mean age of 36.9 years (SD=11.5). Volunteers in excellent health were personnel, relatives of personnel, staff and coworkers, and acquaintances of MDD patients. Both cases and controls underwent identical assessment procedures and were interviewed using the same tools by the same investigator. MDD was identified utilizing the Mini International Neuropsychiatric Interview (M.I.N.I.), translated into Thai by [34], alongside the DSM-5 criteria. The diagnosis of MetS was established utilizing the criteria set out by the American Heart Association/National Heart, Lung, and Blood Institute [35].

The subsequent exclusion criteria pertain to both patients and controls: i) disorders falling under DSM-TR axis-2, including borderline personality disorder and antisocial personality disorder; ii) psychiatric disorders other than MDD and classified as axis-1, encompassing bipolar disorder; psycho-organic syndromes; schizophrenia; substance use disorders (excluding tobacco use disorder), autism spectrum disorders, and schizo-affective psychosis; iii) neurological, neuroinflammatory and neurodegenerative disorders including Alzheimer’s and Parkinson’s disease, stroke, multiple sclerosis, brain tumors, amyotrophic lateral sclerosis, diabetes type 2, and epilepsy; iv) systemic autoimmune and immune disorders including psoriasis, systemic lupus erythematosus, inflammatory bowel disease, COPD, rheumatoid arthritis, infections; and heart disease; v) liver disorders and chronic kidney disease; vi) pregnant or lactating women; vii) allergic or inflammatory reactions the last three weeks prior to the study; and viii) use of immunomodulatory medications (lifelong) and therapeutical doses of antioxidants and omega-3 polyunsaturated fatty acids (one month before the study). Moreover, people who satisfied the inclusion criteria for the control group and possessed a positive familial history of mood disorders, suicide, substance use disorders, or experienced current and/or lifetime MDD and dysthymia were excluded from the study.

#### Clinical assessments

MetS was defined according to the 2009 Joint Scientific Statement of the American Heart Association/National Heart, Lung, and Blood Institute [35] as the occurrence of three or more of the following components: a) HDL cholesterol levels below 40 mg/dL in men and below 50 mg/dL in women; b) triglyceride levels at 150 mg/dL; c) waist circumference of at least 90 cm in men and 80 cm in women; d) fasting blood glucose (FBG) levels elevated to 100 mg/dL or a diabetes diagnosis; e) blood pressure exceeding 130 mm Hg systolic or 85 mm Hg diastolic. The body mass index (BMI) was computed using the formula weight (kg) divided by height (m) squared. The waist circumference was measured at the midpoint between the lowest rib and the superior aspect of the hipbone. The evaluation of ACEs was performed using a Thai version of the ACE Questionnaire [36]. For this research, the raw total scores across several subdomains were calculated and aggregated. Dysthymia and the anxiety disorders outlined in the DSM-IV-TR (post-traumatic stress disorder, panic disorder, and generalized anxiety disorder) were diagnosed utilizing the Mini International Neuropsychiatric Interview (M.I.N.I.) [34] and the DSM-IV-TR criteria. The diagnostic criteria for TUD were applied as specified in the DSM-5.

The Thai translation of the Big Five Inventory (BFI) [37, 38] was utilized to evaluate neuroticism scores. The Columbia Suicide Severity Rating Scale (C-SSRS) [39] was employed to evaluate the severity of both past (lifetime, prior to the index episode) and present (index episode) suicidal ideation (SI) and suicidal attempts (SA). The C-SSRS was employed to assess the frequency, severity, intensity, and lethality of SI and SA. “Current” (Current_SI or Current_SA) refers to the symptoms observed during the final month of the index episode, while lifetime (LT_SI and LT_SA) pertains to symptoms that manifested previously.

Sociodemographic and clinical data were collected through the administration of a semi-structured questionnaire and interviews with patients and controls. The severity of MDD was assessed utilizing the Beck Depression Inventory II (BDI-II) [40, 41] and the Hamilton Depression rating Scale (HAMD) [42]. We used the Thai adaptation of the State-Trait Anxiety Inventory (STAI) [43, 44] to evaluate self-reported anxiety intensity. The HR-QoL was assessed utilizing the World Health Organization Quality of Life Instrument-Abbreviated version (WHO-QoL-BREF) [45]. This measure evaluates four subdomains: physical health, psychological health, social interactions, and environment. The raw subdomain scores were calculated, and subsequently, all scores were aggregated to serve as an index of total HR-QoL.

This study (Electronic Supplementary File or ESF, Table 1) employs clinical composite scores that evaluate i) lifetime pathology of depression-related disorders as a z composite score derived from neuroticism, dysthymia, and the number of anxiety disorders (DYSNEUANX); ii) recurrence of illness (ROI), based on assessments of illness recurrence and suicidal behaviors (SBs); iii) current phenome severity assessed through HAMD, BDI, and STAI scores (z current phenome); iv) total suicidal behaviors (SBs) defined as the aggregate of current and lifetime suicidal ideation and attempts; v) overall severity of depression (OSOD), conceptualized as a composite encompassing all depression features [46].

**Table 1.**
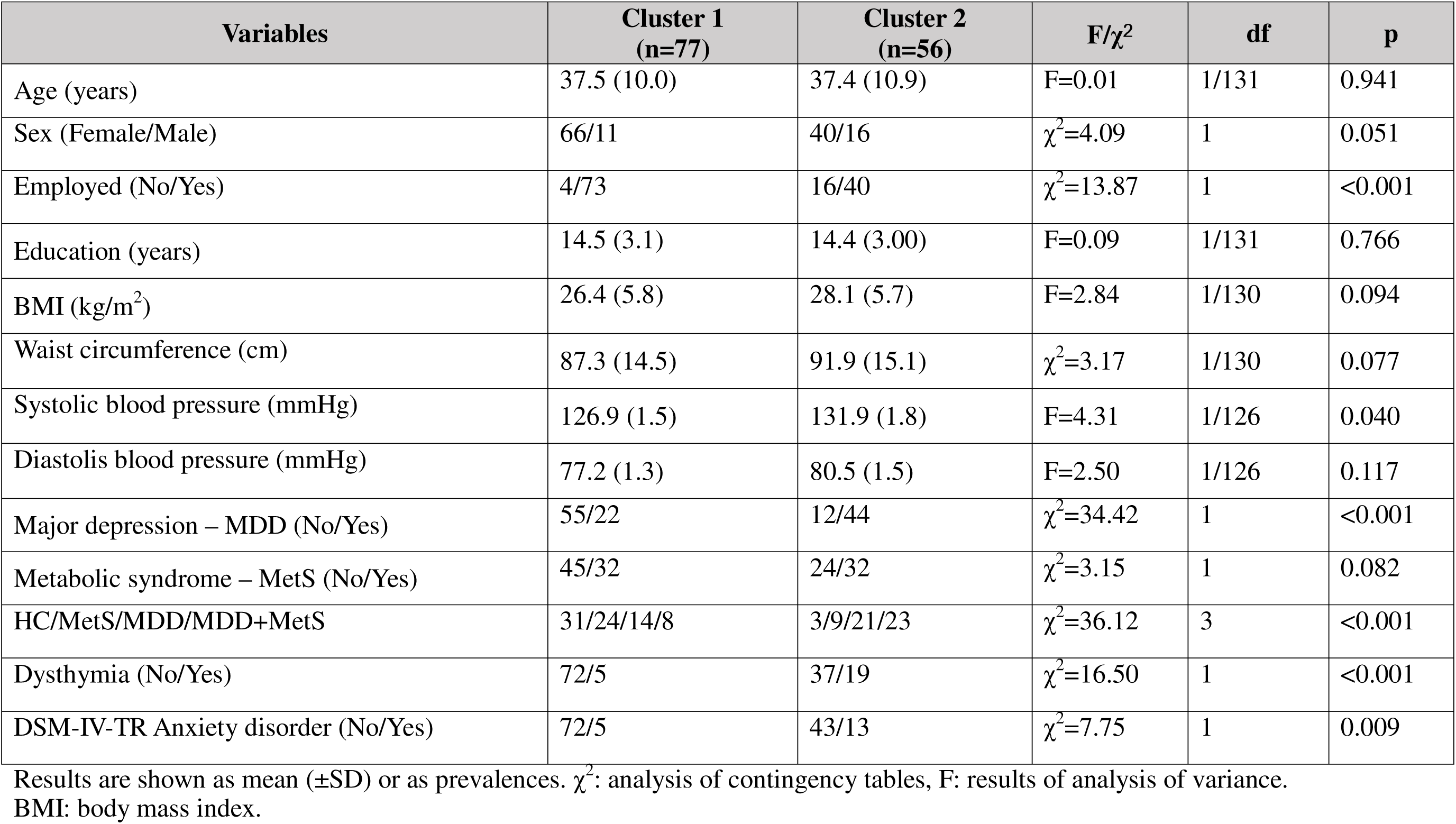
Demographic and clinical data of the study participants divided into two groups using cluster analysis.

| Variables | Cluster 1<br>(n=77) | Cluster 2<br>(n=56) | F/$\chi^2$ | df | p |
| --- | --- | --- | --- | --- | --- |
| Age (years) | 37.5 (10.0) | 37.4 (10.9) | F=0.01 | 1/131 | 0.941 |
| Sex (Female/Male) | 66/11 | 40/16 | $\chi^2=4.09$ | 1 | 0.051 |
| Employed (No/Yes) | 4/73 | 16/40 | $\chi^2=13.87$ | 1 | <0.001 |
| Education (years) | 14.5 (3.1) | 14.4 (3.00) | F=0.09 | 1/131 | 0.766 |
| BMI (kg/m<sup>2</sup>) | 26.4 (5.8) | 28.1 (5.7) | F=2.84 | 1/130 | 0.094 |
| Waist circumference (cm) | 87.3 (14.5) | 91.9 (15.1) | $\chi^2=3.17$ | 1/130 | 0.077 |
| Systolic blood pressure (mmHg) | 126.9 (1.5) | 131.9 (1.8) | F=4.31 | 1/126 | 0.040 |
| Diastolis blood pressure (mmHg) | 77.2 (1.3) | 80.5 (1.5) | F=2.50 | 1/126 | 0.117 |
| Major depression – MDD (No/Yes) | 55/22 | 12/44 | $\chi^2=34.42$ | 1 | <0.001 |
| Metabolic syndrome – MetS (No/Yes) | 45/32 | 24/32 | $\chi^2=3.15$ | 1 | 0.082 |
| HC/MetS/MDD/MDD+MetS | 31/24/14/8 | 3/9/21/23 | $\chi^2=36.12$ | 3 | <0.001 |
| Dysthymia (No/Yes) | 72/5 | 37/19 | $\chi^2=16.50$ | 1 | <0.001 |
| DSM-IV-TR Anxiety disorder (No/Yes) | 72/5 | 43/13 | $\chi^2=7.75$ | 1 | 0.009 |

Results are shown as mean ( ± SD) or as prevalences. $\chi^2$ : analysis of contingency tables, F: results of analysis of variance. BMI: body mass index.

#### Assays

From 8:00 to 9:00 a.m., we collected twenty-five milliliters of fasting venous blood from each study participant utilizing a serum tube and a disposable syringe for the evaluation of cytokines/chemokines/growth factors, lipids, oxidative stress markers, FBG and insulin. Subsequently, the blood was subjected to centrifugation at 3500 revolutions per minute, yielding serum that was preserved in Eppendorf containers with aliquots at -80 degrees Celsius until thawed for biomarker testing. Forty-eight cytokines/chemokines/growth factors/colony stimulating factors were evaluated for their fluorescence intensities (FI) using the Bio-Plex Multiplex Immunoassay reagent provided by Bio-Rad Laboratories Inc., Hercules, USA. A detailed account of our methodologies has been previously published [32]. This study incorporated two principal component scores derived from differentially expressed proteins (DEPs): IM-NT derived from upregulated differently expressed proteins (DEPs) associated with depression and IM-NP derived from downregulated DEPs [32]. Additionally, this study incorporates evaluations of IRS, CIRS, and the z IRS-z CIRS ratio (see to ESF, Table 3) [32, 47]. The electronic supplementary file (ESF), Table 2, contains a list of the other analytes utilized in our study. In this work, we assessed additional variables AOPP, NOx, PON1 Q192R genetic variants, the activities of both catalytic sites of the PON1 enzyme, namely chloromethyl phenylacetate (CMPA)-ase and arylesterase (AREase), FBG and insulin [22–51]. Furthermore, we calculated the ratio of CMPAase to AREase activities (z CMPAase - z AREase), reflecting the difference in activity between both catalytic sites [49]. ESF, Table 2 delineates the methodologies and coefficients of variance for the various lipid fractions assessed in this investigation. The esterified cholesterol ratio, indicative of LCAT activity, was calculated using the method (1 - free cholesterol / total cholesterol) × 100 [7]. We employed four distinct atherogenicity indicators (ESF, table 1): the ApoB/ApoA ratio, Castelli risk index 1, the AIP, and a comprehensive atherogenicity index (CAI) [5, 6, 14, 20, 46]. Additionally, an RCT index (referred to as RCT) was calculated as follows: z HDL-cholesterol + z LCAT + z ApoA1 + z PON1 CMPAase [5]. Consequently, we calculated the z CAI / z RCT ratio as a comprehensive measure of atherogenicity in relation to atheroprotection. ESF, table 2 describes the assays of FBG and insulin. Subsequently, we computed an index of insulin resistance as z glucose – z insulin [28].

**Table 2.** Clinical scores in the cluster analysis-generated classes.

| **Variables** | **Cluster 1**<br>**HC (n=77)** | **Cluster 2**<br>**(n=56)** | **F** | **df** | **p** |
| --- | --- | --- | --- | --- | --- |
| Total adverse childhood experiences score | 1.48 (1.85) | 2.75 (2.30) | 12.41 | 1/131 | <0.001 |
| Neuroticism score | 20.1 (6.4) | 26.4 (6.7) | 30.42 | 1/131 | <0.001 |
| Dysthymia + Neuroticism + Anxiety (z score) | -0.387 (0.626) | 0.520 (1.168) | 32.66 | 1/131 | <0.001 |
| Recurrence of illness (z score) | -0.391 (0.673) | 0.538 (1.116) | 35.62 | 1/131 | <0.001 |
| Beck Depression Inventory (BDI) | 9.7 (9.8) | 25.0 (15.7) | 24.26 | 1/131 | <0.001 |
| Hamilton Anxiety Rating Scale (HAMD) | 6.0 (7.1) | 15.8 (9.2) | 48.44 | 1/131 | <0.001 |
| State and Trait Anxiety Inventory, state version (STAI) | 40.5 (9.7) | 51.3 (10.8) | 36.80 | 1/131 | <0.001 |
| Current BDI + HAMD + STAI (z score) | -0.457 (0.720) | 0.628 (0.986) | 53.86 | 1/131 | <0.001 |
| Total suicidal behaviors (z score) | -0.334 (0.561) | 0.468 (1.258) | 24.57 | 1/130 | <0.001 |
| Overall severity of depression (z score) | -0.447 (0.685) | 0.625 (1.031) | 51.42 | 1/130 | <0.001 |
| Total Health-Related Quality of Life | 88.4 (12.8) | 72.6 (16.3) | 39.22 | 1/131 | <0.001 |

Results are shown as mean ( ± SD). F: results of analysis of variance.

**Table 3.** Biomarker measurements in the cluster-analysis generated classes.

| **Variables** | **Cluster 1**<br>**HC (n=77)** | **Cluster 2**<br>**(n=56)** | **F/$\chi^2$** | **df** | **p** | **FDR p**<br>**values** |
| --- | --- | --- | --- | --- | --- | --- |
| Paraoxonase (PON)1 genotypes (Q/Q, Q/R, R/R) | 11 / 22 / 44 | 10 / 25 / 21 | 5.19 | 2 | 0.083 | 0.115 |
| PON1 R/R 192 (No/Yes) | 33 / 44 | 26 / 21 | 5.01 | 1 | 0.035 | 0.051 |
| PON1 chloromethyl phenylacetate (CMPA)ase (U/mL) | 45.45 (2.08) | 38.76 (2.63) | 4.45 | 1/161 | 0.002 | 0.004 |
| PON1 arylesterase (AREase) (U/mL) | 235.6 (9.5) | 249.6 (9.46) | 0.37 | 1/131 | 0.614 | 0.685 |
| PON1 z CMPAase – z AREase (z score) | 0.261 (0.142) | -0.359 (0.166) | 8.03 | 1/131 | 0.005 | 0.010 |
| Advanced oxidation protein products (μmol/L/eq. chloramine T) | 123.94 (16.87) | 216.05 (19.82) | 24.07 | 1/126 | <0.001 | 0.002 |
| Nitric oxide metabolites (μmol/L) | 6789 (391) | 6576 (466) | 0.12 | 1/123 | 0.731 | 0.757 |
| Immune-linked neurotoxicity (IM-NT) (z scores) | -0.423 (0.101) | 0.566 (0.119) | 39.01 | 1/126 | <0.001 | 0.002 |
| Immune-linked neuroprotection (IM-NP) (z scores) | 0.042 (0.118) | -0.044 (0.138) | 0.22 | 1/126 | 0.641 | 0.688 |
| IRS (z scores) | -0.268 (0.109) | 0.353 (0.129) | 13.17 | 1/126 | <0.001 | 0.002 |
| CIRS (z scores) | -0.111 (0.113) | 0.133 (0.133) | 1.90 | 1/126 | 0.171 | 0.216 |
| z IRS - z CIRS (z scores) | -0.185 (0.115) | 0.259 (0.135) | 6.11 | 1/126 | 0.015 | 0.024 |
| TNF-signaling (z scores) | -0.307 (0.108) | 0.409 (0.127) | 18.03 | 1/126 | <0.001 | 0.002 |
| Insulin (mIU/L) | 12.98 (0.99) | 14.34 (1.17) | 0.796 | 1/126 | 0.374 | 0.452 |
| Fasting Blood Glucose (FBG) (mg/dL) | 96.33 (3.08) | 94.90 (3.62) | 0.089 | 1/126 | 0.766 | 0.766 |
| z FBG – z Insulin (z score) | 0.040 (0.140) | -0.301 (0.167) | 0.101 | 1/119 | 0.403 | 0.452 |
| Apolipoprotein E (ApoE) (mg/dL) | 931.9 (44.8) | 1192.2 (52.7) | 13.82 | 1/126 | <0.001 | 0.002 |
| Free cholesterol (mg/dL) | 60.9 (2.4) | 69.7 (2.9) | 5.33 | 1/126 | 0.023 | 0.035 |
| HDL cholesterol (mg/dL) | 57.54 (1.27) | 49.48 (1.49) | 16.42 | 1/126 | <0.001 | 0.002 |
| ApoA (mg/dL) | 145.2 (2.9) | 129.8 (3.5) | 11.20 | 1/126 | 0.001 | 0.002 |
| Esterified cholesterol ratio (LCAT) % | 70.0 (1.0) | 65.7 (1.2) | 6.49 | 1/126 | 0.010 | 0.017 |
| Reverse cholesterol transport (RCT) (z score) | 0.294 (0.098) | -0.424 (0.115) | 22.14 | 1/126 | <0.001 | 0.002 |
| ApoB (mg/dL) | 93.7 (2.8) | 100.7 (3.3) | 2.53 | 1/126 | 0.114 | 0.150 |
| Triglycerides (mg/dL) | 122.9 (19.1) | 206.6 (22.4) | 7.98 | 1/126 | 0.006 | 0.011 |
| Comprehensive atherogenicity index (CAI) (z score) | -0.202 (0.093) | 0.317 (0.109) | 12.72 | 1/126 | 0.001 | 0.002 |
| z CAI – z RCT (z score) | -0.496 (0.141) | 0.741 (0.165) | 31.59 | 1/126 | <0.001 | 0.002 |
| ApoB / ApoA | 0.677 (0.025) | 0.796 (0.029) | 9.27 | 1/126 | 0.003 | 0.006 |
| Castelli Risk Index 1 (total cholesterol / HDL) | 3.73 (0.12) | 4.37 (0.14) | 12.33 | 1/126 | 0.001 | 0.002 |
| Triglycerides / HDL (Atherogenicity index of plasma) | 2.33 (0.23) | 3.88 (0.23) | 26.57 | 1/126 | <0.001 | 0.002 |

All results (except the PON1 data) are shown as estimated marginal means (SE) after covarying for age, sex, body mass index, waist circumference, and metabolic syndrome. FDR: false discovery rate p correction.
IRS: immune-inflammatory response system; CIRS: compensatory immunoregulatory system; TNF: tumor necrosis factor; HDL: high-density lipoprotein; z CMPAase – z AREase: ratio of the two catalytic PON1 sites.

#### Statistical analysis

The chi-square test or Fisher’s Exact Probability Test was employed to compare nominal variables across different categories. Analysis of variance was utilized to examine scale variables across several diagnostic groups. The false discovery rate (FDR) p-correction was applied to the multiple comparisons of biomarker data. The data distributions were subjected to essential normalizing procedures, encompassing rank-based, logarithmic, square-root, and inverse normal transformations. Furthermore, z-score modification was utilized on the data to enhance interpretability and provide z-unit-weighted composite scores that more accurately depict various biomarker profiles. Pearson’s product moment coefficients were utilized to ascertain the correlations between two sets of scale variables. Multiple regression analysis was employed to evaluate the impact of immunological, oxidative stress and insulin indicators on lipid variables, while allowing for the effects of age, sex, MetS, BMI, and waist circumference. We employed both manual and automatic forward stepwise regression methods, utilizing p-values of 0.05 for inclusion and 0.06 for exclusion. The final regression models incorporated standardized coefficients, t-statistics, and exact p-values for each explanatory variable, in addition to total variance explained (shown by R^2^ or partial Eta squared as effect size) and F statistics (together with p values). Collinearity and multicollinearity were assessed using tolerance (cut-off value <0.25), variance inflation factor (cut-off value >4), condition index, and variance proportions from the collinearity diagnostics table. The White and modified Breusch-Pagan tests were employed to ascertain the presence of heteroskedasticity. Two-step cluster analysis was conducted to identify subclasses of individuals based on present phenome severity and NIMETOX variables. The approach employed the log-likelihood distance metric and the Schwarz Bayesian clustering criterion. The cluster solution was deemed acceptable when the silhouette measure of cohesion and separation reached a minimum of 0.4 and the ratio of the largest to smallest cluster did not exceed 2. A binary logistic regression analysis was performed to identify the primary predictors of the established clusters by incorporating variables that were not utilized in the cluster formation (e.g., HR-QoL, PON1 activity, ROI, etc.). We utilized both manual and automatic forward stepwise regression methods, applying p-values of 0.05 for inclusion and 0.06 for exclusion. We calculated odds ratios with 95% confidence intervals, B (SE) and Wald (df and p) statistics for the significant variables in the final model, along with Nagelkerke pseudo R-squared (used as effect size) and accuracy (including sensitivity and specificity). A two-tailed design was utilized in each of the previously listed studies, with statistical significance established at an alpha level of 0.05. The program employed was IBM Windows SPSS version 28. Principal component analysis (PCA) was implemented to extract PCs from the biomarker data set and varimax rotation was implemented to interpret the PCs when more than one PC could be extracted. The PC model was only accepted when the Kaiser-Meyer-Olkin (KMO) measure of sampling adequacy was greater than 0.7 and the Bartlett’s test of sphericity was significant. The number of PCs was determined by a 50% explained variance. The only permissible loadings on the PCs were those that exceeded 0.4. As a result, PC scores were computed and implemented in additional statistical analyses.

The *a priori* sample size was calculated using G*Power version 3.1.9.4. The primary statistical analysis conducted was a multiple regression analysis, using the z CAI – z RCT score as the dependent variable and NIMETOX data as independent factors. With f = 0.136 (representing around 12% of the explained variance), five explanatory variables, an alpha level of 0.05, and a power of 0.8, the calculated minimum sample size was 100. Given the potential disparities in correlations between persons with and without MetS, we conducted exploratory analyses for both groups.

## Results

### Cluster analysis

Cluster analysis performed on the current phenome, AOPP, AIP and IM-NT generated a two-cluster solution with a silhouette measure of cohesion and separation of 0.5 (average). The largest cluster (cluster 1) contained 77 individuals (57.9%) and the smallest cluster (cluster 2) 56 individuals (42.1%). The largest/smallest cluster ratio was 1.38. **Table 1** shows the demographic and clinical data of the study participants divided into these two clusters. There were no significant differences in age, sex ratio, education years, BMI, waist circumference, diastolic blood pressure, and MetS between these two clusters. Unemployment, systolic blood pressure, and the prevalence of MDD, dysthymia and anxiety disorders were significantly higher in cluster 2 than in cluster 1.

### Depression features differences between both clusters

**Table 2** shows the differences in rating scale scores between participants divided into two clusters. Those allocated to cluster 2 showed significantly higher scores on total ACEs, neuroticism, DYSNEUANX, ROI, current BDI, HAMD and STAI, current phenome, total suicidal behaviors, and OSOD. In addition, cluster 2 was characterized by lower HR-QoL scores as compared with cluster 1.

### Biomarker differences between both clusters

**Table 3** shows the biomarker measurements in the study participants divided into the cluster analysis-generated groups. Without p-corrections the RR PON1 genotype was more prevalent in cluster 2 than cluster 1 and PON1 CMPAase activity and the zCMPAase – z AREase score were significantly lower in cluster 2 than cluster 1. AOPP, but not NOx, levels were significantly increased in cluster 2 compared to cluster 1. IM-NT, IRS, z IRS – z CIRS, and TNF-signaling scores were significantly higher in cluster 2 compared to cluster 1. There were no significant differences in CIRS values between both groups. There were no significant differences in insulin, FBG or z FBG – z insulin, between the two study groups. ApoE, free cholesterol, triglycerides, CAI, and z CAI-z RCT were significantly higher in cluster 2 than cluster 1. HDL cholesterol, ApoA, LCAT, and RCT were significantly lower in cluster 2 than in cluster 1. The three classical atherogenicity indices used in this study (ApoB/ApoA; Castelli risk index 1, and AIP) were significantly higher in cluster 2 than cluster 1. Since cluster 2 was characterized by increased NIMETOX biomarkers, it was denoted as the NIMETOX cluster.

Three normal controls (no MDD and no MetS present) and 9 patients with MetS (without MDD) were allocated to the NIMETOX cluster. All three normal controls showed very high IM-NT and very high TNF signaling, very low CMPAase, and one of the controls showed very low LCAT and RCT coupled with increased atherogenicity. The ApoB/A ratio was significantly increased in two of these controls. The nine MetS subjects allocated to the NIMETOX cluster showed very low CMPAase activities, RCT and LCAT activity (n=6), increased AOPP (n=4) and atherogenicity indices, including high Castelli risk index 1 indices (all nine), while CAI was increased in eight of them.

**Table 4** shows the outcome of a binary logistic regression analysis with the NIMETOX cluster as dependent variable and clinical data or biomarkers that are not used to generate the cluster as input variables. We found that cluster 2 was characterized by increased ROI, and lowered PON1 CMPAase activity and HR-QoL with an accuracy of 78.8% (sensitivity=69.1% and specificity=85.7%) and Nagelkerle effect size of 0.486 (χ^2^=59.06, df=3, p<0.001).

**Table 4.** Results of logistic regression analyses with the NIMETOX cluster (cluster 2) as dependent variable.

| **Dependent variable** | **Explanatory variables** | **B** | **SE** | **W** | **df** | **p** | **OR** | **95% CI**<br>**Lower / upper** |
| --- | --- | --- | --- | --- | --- | --- | --- | --- |
| Cluster 2 vs<br>cluster 1 | ROI | 1.044 | 0.306 | 11.67 | 1 | <0.001 | 2.841 | 1.560 / 5.171 |
|  | PON1 CMPAase | -0.912 | 0.253 | 16.05 | 1 | <0.001 | 0.402 | 0.245 / 0.659 |
|  | HR-QoL | -0.60 | 0.017 | 12.85 | 1 | <0.001 | 0.942 | 0.912 / 0.973 |

ROI: recurrence of illness index; PON1: paraoxonase 1; CMPAase chloromethyl phenylacetate (CMPA)-ase; HR-QoL; total health-related quality of life score.

### No effects of medical treatments

Previously, we have reported that there are no significant effects of the drug state of the subjects on the LCAT, RCT, and atherogenity biomarkers, and the immune biomarkers measured in this study population [14, 32]. Some of the participants were administered antidepressants (n=62), benzodiazepines (n=56), antipychotic drugs (n=14), simvastatin (n=19), metformin (n=11), and all metabolic medications combined (n=31, including lipid lowering, antihypertensive and anti-diabetic medications). In the present study, there were no significant effects of antidepressants (p=0.941), benzodiazepines (p=0.434), antipsychotic drugs (p=0.989), simvastatin (p=0.612), metformin (p=0.594) and all metabolic medications combined (p=0.543) on FBG, insulin and the insulin resistance index. There were no significant effects of antidepressants (p=0.591), benzodiazepines (p=0.838), antipsychotic drugs (p=0.959), simvastatin (p=0.570), metformin (p=0.416) and all metabolic medications combined (p=0.415) on AOPP, and PON1 CMPAase and AREase activities. Therefore, there is no evidence that the results are influenced by the drug state.

### Intercorrelations between the lipid and other biomarkers

**Table 5** lists the outcome of different multiple regression analyses with the lipid biomarkers as output variables and immune, oxidative stress and insulin resistance biomarkers as input variables. Regression #1 shows that 44.5% of the variance in the z CAI – z RCT index was associated with FBG, AOPP, male sex, and IRS (all positive associated) and PON1 AREase activity (inversely associated). Regression #2 shows that 26.1% of the variance in RCT was associated with IRS and FBG (both inversely) and the PON1 genotype additive model (entered as QQ=0, QR=1, RR=2). A large part of the variance in CAI (regression #3) was explained by AOPP, FBG, male sex and age (all positively associated). 25.4% of the variance in ApoA (regression #4) was associated with male sex (inversely associated) and PON1 CMPAase and age (positively associated). A large part of the variance in HDL was explained by FBG, insulin, male sex, and IRS (all inversely associated) and PON1 CMPAase (positively associated). We found that 18.3% of the variance in the LCAT index was explained by IRS, FBG, AOPP (all three negatively associated) and PON1 CMPAase (positively associated). Male sex, FBG, AOPP (positively associated) and PON1 CMPAase (inversely associated) explained 42.6% of the variance in the Castelli risk index 1.

**Table 5.** Results of multiple regression analysis with atherogenicity-related biomarkers as dependent variables.

| Dependent Variables | Explanatory Variables | Coefficients of input variables |  |  | Model statistics |  |  |  |
| --- | --- | --- | --- | --- | --- | --- | --- | --- |
| | | $\beta$ | t | p | R<sup>2</sup> | F | df | p |
| #1. CAI / RCT | **Model #1** |  |  |  | 0.445 | 20.33 | 5/127 | <0.001 |
|  | FBG | 0.354 | 5.23 | <0.001 |  |  |  |  |
|  | AOPP | 0.292 | 4.29 | <0.001 |  |  |  |  |
|  | PON1 AREase | -0.307 | -4.56 | <0.001 |  |  |  |  |
|  | Sex | 0.201 | 2.94 | 0.004 |  |  |  |  |
|  | IRS | 0.189 | 2.83 | 0.005 |  |  |  |  |
| #2. RCT | **Model #2** |  |  |  | 0.261 | 14.82 | 3/126 | <0.001 |
|  | IRS | -0.338 | -4.41 | <0.001 |  |  |  |  |
|  | FBG | -0.323 | -4.22 | <0.001 |  |  |  |  |
|  | PON1 Q192R additive model | 0.224 | 2.92 | 0.004 |  |  |  |  |
| #3. CAI Atherogenicity | **Model #3** |  |  |  | 0.446 | 27.74 | 4/128 | <0.001 |
|  | AOPP | 0.466 | 6.98 | <0.001 |  |  |  |  |
|  | FBG | 0.296 | 4.22 | <0.001 |  |  |  |  |
|  | Sex | 0.229 | 3.36 | 0.001 |  |  |  |  |
|  | Age | 0.159 | 2.29 | 0.023 |  |  |  |  |
| #4. ApoA | **Model #4** |  |  |  | 0.254 | 14.47 | 3/129 | <0.001 |
|  | PON1 CMPAase | 0.390 | 5.01 | <0.001 |  |  |  |  |
|  | Sex | -0.341 | -4.45 | <0.001 |  |  |  |  |
|  | Age | 0.199 | 2.27 | 0.011 |  |  |  |  |
| **#5. HDL** | **Model #5** |  |  |  | 0.363 | 14.47 | 5/127 | <0.001 |
|  | FBG | -0.319 | -4.34 | <0.001 |  |  |  |  |
|  | Sex | -0.312 | -4.22 | <0.001 |  |  |  |  |
|  | PON1 CMPAase | 0.243 | 3.32 | 0.001 |  |  |  |  |
|  | Insulin | -0.185 | -2.53 | 0.013 |  |  |  |  |
|  | IRS | -0.150 | -2.07 | 0.040 |  |  |  |  |
| **#6. LCAT** | **Model \$6** | | | | 0.183 | 6.99 | 4/125 | <0.001 |
|  | IRS | -0.205 | -2.47 | 0.015 |  |  |  |  |
|  | FBG | -0.232 | -2.86 | 0.005 |  |  |  |  |
|  | AOPP | -0.198 | -2.41 | 0.017 |  |  |  |  |
|  | PON1 CMPAase | 0.173 | 2.05 | 0.042 |  |  |  |  |
| **#7. Castelli risk index 1** | **Model #7** |  |  |  | 0.426 | 23.80 | 4/128 | <0.001 |
|  | Sex | 0.466 | 6.72 | <0.001 |  |  |  |  |
|  | FBG | 0.258 | 3.78 | <0.001 |  |  |  |  |
|  | AOPP | 0.248 | 3.60 | <0.001 |  |  |  |  |
|  | PON1 CMPAase | -0.209 | -3.06 | 0.003 |  |  |  |  |

CAI: comprehensive atherogenicity index; RCT: reverse cholesterol transport; FBG: fasting blood glucose; AOPP: advanced oxidation protein products; PON1: paraoxonase 1; AREase: arylesterase; CMPAase chloromethyl phenylacetate (CMPA)-ase; Apo: apolipoprotein; HDL: high-density lipoprotein; LCAT: lecithin-cholesterol acyltransferase; Sex: male=1, female=0; IRS: immune-inflammatory responses system; Castelli 1: total cholesterol/HDL.

### Association with depression features

**Table 6** lists the results of multiple regression analysis with the clinical composites as dependent variables and all biomarkers and the total ACE score as explanatory variables, while allowing for the effects of age, sex, and education. Regression #1 shows that 42.1% of the variance in neuroticism was explained by the total ACE score, IM-NT, insulin (all positively correlated) and IM-NP (inversely correlated). We have also examined the same regression but now in people with versus no MetS and found quite different predictors in both study groups. Thus, in individuals without MetS, we found that ACEs (p<0.001), ApoB/ApoA (p=0.006) (both positively) and PON1 AREase (p=0.048) (inversely) explained 32.3% of the variance (F=10.36, df=3/65, p<0.001). In those with MetS a quite different model was found, namely 42.2% of the variance was explained (F=14.63, df=3/60, p<0.001) by IM-NT (p=0.002), insulin (p<0.001), and ACEs (p=0.001) (all three positively).

**Table 6.** Results of multiple regression analysis with clinical composite scores as dependent variables.

| Dependent Variables | Explanatory Variables | Coefficients of input variables |  |  | Model statistics |  |  |  |
| --- | --- | --- | --- | --- | --- | --- | --- | --- |
| | | $\beta$ | t | p | $R^2$ | F | df | p |
| #1. Neuroticism | **Model #1** |  |  |  | 0.421 | 23.27 | 4/128 | <0.001 |
|  | ACE | 0.465 | 6.74 | <0.001 |  |  |  |  |
|  | IM-NP | -0.318 | -4.58 | <0.001 |  |  |  |  |
|  | IM-NT | 0.245 | 3.53 | 0.001 |  |  |  |  |
|  | Insulin | 0.168 | 2.49 | 0.014 |  |  |  |  |
| #2. Dysthymia +<br>Neuroticism +<br>Anxiety (z score) | **Model #2** |  |  |  | 0.311 | 11.49 | 5/127 | <0.001 |
|  | ACE | 0.362 | 4.79 | <0.001 |  |  |  |  |
|  | IM-NT | 0.239 | 3.12 | 0.002 |  |  |  |  |
|  | IM-NP | -0.220 | -2.90 | 0.004 |  |  |  |  |
|  | ApoB/ApoA | 0.174 | 2.35 | 0.020 |  |  |  |  |
|  | Insulin | 0.160 | 2.16 | 0.033 |  |  |  |  |
| #3. Current phenome | **Model #3** |  |  |  | 0.520 | 27.48 | 5/127 | <0.001 |
|  | ACE | 0.491 | 7.79 | <0.001 |  |  |  |  |
|  | IM-NP | -0.349 | -5.50 | <0.001 |  |  |  |  |
|  | AOPP | 0.200 | 3.21 | 0.002 |  |  |  |  |
|  | Insulin | 0.211 | 3.38 | 0.001 |  |  |  |  |
|  | IM-NT | 0.212 | 3.33 | 0.002 |  |  |  |  |
| #4. LT SB | **Model #4** |  |  |  | 0.277 | 12.15 | 4/127 | <0.001 |
|  | ACE | 0.383 | 5.03 | <0.001 |  |  |  |  |
|  | AOPP | 0.203 | 2.67 | 0.009 |  |  |  |  |
|  | ApoB/ApoA | 0.166 | 2.17 | 0.032 |  |  |  |  |
|  | IM-NT | 0.160 | 2.08 | 0.039 |  |  |  |  |
| **#4. Overall severity of depression (OSOD)** | **Model #5** |  |  |  | 0.504 | 21.15 | 6/125 | <0.001 |
|  | ACE | 0.482 | 7.45 | <0.001 |  |  |  |  |
|  | AOPP | 0.177 | 2.74 | 0.007 |  |  |  |  |
|  | IM-NT | 0.270 | 4.12 | <0.001 |  |  |  |  |
|  | IM-NP | -0.280 | -4.28 | <0.001 |  |  |  |  |
|  | Insulin | 0.217 | 3.38 | 0.001 |  |  |  |  |
|  | Sex | 0.152 | 2.33 | 0.021 |  |  |  |  |

ACE: adverse childhood experiences; IM-NP: immune-linked neuroprotection; IM-NT: immune-linked neurotoxicity; AOPP: advanced oxidation protein products; Apo: apolipoprotein; sex: male=1, female=0.

Table 6, regression #2 shows that 31.1% of the variance in the DYSNEUANX index was associated with IM-NP (inversely) and 4 positively associated predictors, namely ACEs, IM-NT, ApoB/ApoA, and insulin. There were again major differences between people with and without MetS. In those with MetS, 20.1% of the variance in this index was explained by IM-NT (p<0.001), whereas in those without MetS, 37.4% of the variance was explained by AIP (p<0.001), ACEs (p=0.011) and NOx (p=0.019).

Regression #3 shows that 52.0% of the variance in the current phenome was explained by the regression on ACEs, AOPP, insulin, IM-NT (all four positively associated) and IM-NP (inversely associated). In those with MetS, 16.5% was explained by IM-NT (p<0.001), whereas in those without MetS, we found that the cumulative effects of Castelli risk index 1 (p<0.001) and ACEs (p<0.001) explained 37.9% of the variance.

Regression #4 shows that lifetime + current SBs were significantly predicted (27.7% of the variance) by ACEs, AOPP, ApoB/ApoA and IM-NT (all positively). In subjects without MetS, 37.9% of the variance (F=18.85, df=2/65, p<0.001) was associated with Castelli risk index 1 (p<0.001) and ACEs (p<0.001). In those with MetS, 16.5% of the variance was positively associated with IM-NT (p<0.001).

Regression #5 shows that OSOD was largely predicted (50.4% of the variance) by IM-NP (inversely) and five other predictors, namely ACEs, AOPP, IM-NT, insulin, and male sex. In those without MetS, we found that 48.7% of the variance was explained by ACEs (p<0.001), AIP (p=0.003) and ApoB/ApoA (p=0.038) (all three positively associated). In those with MetS, 31.1% was explained by the additive effects of IM-NT (p=0.001), insulin (p=0.009) and ACEs (p<0.001). Both scores were significantly higher in cluster 2 than in cluster 1 (p<0.001).

### Principal component analysis (PCA)

In order to determine whether we could extract one or more relevant (and meaningful) principal components (PCs) from a data set comprising ACEs, DYSNEUANX, current phenome, IM-NT, TNF-signaling, AOPP, Castelli risk index 1, CAI, RCT, and FBG, we performed PCA. We could retrieve 2 PCs which together explained 51.4% of the variance. Varimax rotation showed that ACEs (loading=0.929), DYSNEUANX (0.772), current phenome (0.821), IM-NT (0.646), and TNF-signaling (0.657) loaded highly on the first PC which explained 25.96% of the variance. The second varimax-rotated PC explained 25.47% of the variance and loaded highly on Castelli risk index 1 (0.911), CAI (0.907), RCT (-0.618), FBG (0.541), and AOPP (0.427). While the second varimax-rotated PC score was significantly higher in those with MetS versus those without (F=38.13, df=1/129, p<0.001) there were no significant differences in the first PC score (F=0.02, df=1/129, p=0.892).

## Discussion

### The NIMETOX Cluster

The primary outcome of this study reveals that we successfully identified a cohort of individuals exhibiting elevated scores on depressive features, alongside nearly all NIMETOX pathways. This includes heightened atherogenicity, diminished LCAT and RCT functions, activation of IM-NT cytokine networks, increased protein oxidation, and reduced antioxidant defenses, as evidenced by decreased PON1 CMPAase activities. It is important to emphasize that insulin resistance did not play a significant role in this NIMETOX cluster.

As indicated by the data, a significant proportion, specifically 78.8%, of patients diagnosed with OMDD were assigned to this newly established NIMETOX cluster. Furthermore, the patients diagnosed with OMDD that were not assigned to the NIMETOX-depression cluster exhibited varying levels of NIMETOX biomarkers, either exceeding or falling below the mean values observed within the non-NIMETOX-depression cohort. This included instances of increased AOPPs in one case, notably low levels of CMPAase in two cases, and heightened atherogenicity as indicated by a high Castelli risk index in two instances, and elevated ApoB/ApoA ratios in two cases. Additionally, there was a significantly reduced RCT activity in four cases and lowered LCAT activity in one case, alongside increased IM-NT in one case and TNF signaling in two cases.

It is important to highlight that the inclusion rate of 78.8% for OMDD patients within the NIMETOX cluster could be enhanced by incorporating additional immune-metabolic biomarkers that exhibit alterations in MDD. These include lipid peroxidation markers such as lipid hydroperoxides (LOOH) and malondialdehyde (MDA) [15, 51], autoimmune responses to oxidized lipids [52], including oxidized LDL, as well as oxidative specific epitopes (OSEs, such as MDA and azelaic acid).

The findings presented here stand in opposition to the assertions that “inflammation in depression” is observed in approximately 30% of MDD cases [53], as well as to the construction of a purported immune-metabolic depression class [54]. The previously cited prevalence rate of 30% for “inflammation in MDD” [53] was derived from an evaluation of increased serum C-reactive protein levels. In contrast, our investigation employed a comprehensive state-of-the art assessment of various biomarkers and pathways to enhance accuracy. Moreover, it is important to note that C-reactive protein levels within the lower secretion ranges do not serve as a reliable indicator of “inflammation,” given that approximately 50% of the variance in the low C-reactive protein range can be attributed to factors such as BMI, ACEs, and age [55]. Furthermore, it is important to note that MDD does not constitute an “inflammatory” disorder [12]. Indeed, the immune disorders associated with MDD are characterized by imbalances within immune profiles, which may also vary depending on the stage of the disorder [9, 10, 11, 12, 13, 14, 22, 32, 46]. One may observe entirely distinct profiles contingent upon the phenotype of MDD, e.g., MDMD (increased IRS versus CIRS) versus SDMD (lowered CIRS) [12, 13, 56]. The present investigation conducted among patients with OMDD elucidates that varying immune profiles correlate with the phenome of depression, characterized by elevated IM-NT levels, diminished IM-NP levels, and heightened TNF-signaling activity. Therefore, papers claiming that the incidence of “inflammation in MDD” may be estimated at around 30% are inaccurate.

It is of paramount importance to note that Penninx et al. incorporated a limited number of biomarkers characterized by low accuracy, including triglyceride and C-reactive protein levels, within their analysis constructing an immune-metabolic class of patients [54]. Consequently, they were unable to investigate the state-of-the-art NIMETOX biomarkers implicated in MDD, some of which had already been identified in the 1990s. The authors in question omitted LCAT activity [7], RCT [5], atherogenicity indices [6, 7], protein oxidation biomarkers such as AOPP [51], and PON1 activity [22], which serves as a significant lipid-protecting enzyme and a crucial element within the NIMETOX cluster (refer to the results of the binary logistic regression analysis), as well as recurrent depression [22]. In addition, the authors in question did not utilize the advanced immune profiling method developed by Maes et al. [12, 56] based on Multiplexing assays, and the construction of relevant immune composites and PCs. Subsequently, these papers are disseminated as a review; however, all primary data and the latest advancements in the field are notably silenced. Furthermore, the findings presented by Penninx et al. [56] pose interpretative challenges, as a) the lipid results were not assessed in individuals both with and without MetS [13, 46]; and b) they did not consider the heterogeneity of depression phenotypes.

### AOPP, immune activation, atherogenicity and OMDD features

In the present investigation, we noted that there exist highly significant correlations among elevated levels of AOPP, immune activation, diminished LCAT activity, heightened atherogenicity, and the severity of phenotypic characteristics associated with OMDD. The results of our investigation build upon earlier research that indicates a correlation between elevated oxidation biomarkers, specifically MDA and AIP as demonstrated by [57].

Furthermore, our study highlights the complex interactions that exist between lipid biomarkers, such as HDL, and immune biomarkers in the context of the acute episode of severe MDD [2, 8]. In OMDD, the association between atherogenicity and elevated levels of reactive oxygen and nitrogen species, as well as oxidative stress, is notably significant, as evidenced by the increased levels of AOPP and MDA [58]. Venturini et al. [59] demonstrated that AOPP exhibited a stronger association with MetS compared to LOOH (lipid hydroperoxides).

The formation of AOPP serves as an indicator of protein oxidation, primarily associated with heightened myeloperoxidase (MPO) activity, which leads to an increased production of hypochlorous acid through the interaction of hydrogen peroxide and chloride ions [21, 51, 60, 61]. Furthermore, AOPP has the potential to exacerbate atherosclerotic lesions, promote the accumulation of macrophages within these lesions, inhibit LCAT activity, and disrupt RCT in murine models [62, 63]. Moreover, AOPP has the potential to disrupt HDL metabolism through the inhibition of the scavenger receptor class B type I (SR-BI) [64]. The latter plays a crucial role in facilitating the cholesterol efflux from macrophages to the HDL particle, a process that significantly contributes to antiatherogenic mechanisms [65, 66].

Prior research has demonstrated a correlation between oxidative stress and heightened atherogenicity as well as insulin resistance in individuals suffering from affective disorders [28]. Elevated levels of oxidative stress have been observed in metabolic syndrome (MetS) and its associated consequences, including coronary artery disease [67]. Atherogenesis represents a multifaceted process that arises not solely from lipid abnormalities but also from immune-inflammatory responses and oxidative stress processes [68]. Indeed, the activation of immune-inflammatory and oxidative pathways serves as a crucial element in the initiation and subsequent stages of atherosclerosis and plaque formation [5, 69].

Conversely, an elevation in atherogenicity may result in heightened oxidative stress. For instance, elevated levels of free fatty acids and FBG contribute to an increase in reactive oxygen species within vascular cells, subsequently leading to the production of proinflammatory cytokines [70]. Furthermore, the cytokines that are found to be elevated in MetS have the potential to induce oxidative stress [70]. In addition, conditions such as hyperglycemia, insulin resistance, and hypertriglyceridemia may contribute to an increase in lipid peroxidation and the formation of aldehydes within the context of MetS [71]. Moreover, infections and conditions marked by immune-inflammatory activation are associated with elevated triglyceride levels and diminished HDL cholesterol levels [72]. In summary, one can observe numerous bidirectional associations among the diverse NIMETOX variables.

### PON1, atherogenicity and OMDD features

Another significant finding of this study is that reduced PON1 CMPAase activity is accompanied by diminished RCT and LCAT activities and is closely linked to heightened atherogenicity and is a highly significant determinant of the NIMETOX cluster. PON1 is an enzyme associated with HDL that serves to protect HDL and LDL from oxidation, thereby exhibiting antiatherogenic, antioxidant and anti-inflammatory properties [22, 23]. The antioxidant characteristics of HDL can be attributed, in part, to the presence of PON1, ApoA, and LCAT molecules that are associated with HDL [5]. For instance, the interplay among Apo A1, LCAT, and PON1 extends the duration during which HDL is capable of inhibiting LDL oxidation, attributable, in part, to the mitigation of LCAT inactivation [73]. These interactions subsequently stimulate the capacity of HDL to facilitate cholesterol efflux [74]. Moreover, PON1 has peroxidase activity thereby neutralizing the detrimental effects of reactive oxygen species and oxygen peroxides [23].

The above findings explain that diminished activities of PON1 CMPAase correlate with heightened atherogenicity [23] and an escalation in disabilities resulting from stroke [48]. It is noteworthy that the present investigation revealed an imbalance between the two catalytic sites of PON1, specifically CMPAase and AREase. This study observed a reduction in CMPAase activity relative to the activity of AREase within the NIMETOX cluster. Prior investigations have indicated that imbalances between the two catalytic sites serve as predictors for the severity of stroke [49].

Moreover, the activities of PON1 constitute an essential component, alongside ApoA and LCAT, within the RCT pathway [5]. Consequently, a reduction in RCT activity correlates not only with lipid irregularities but also with the activation of IRS and an augmented oxidation of HDL and LDL, which is subsequently followed by autoimmune responses to, for instance, oxidized LDL and oxidatively modified lipids [5]. The latter may result in dysfunctional HDL particles that exhibit pro-inflammatory characteristics and potentially facilitate atherogenic processes [5]. In conclusion, diminished PON1 activity, particularly with respect to CMPAase activity, could be instrumental in the cascade of events leading from reduced LCAT and RCT activities, oxidative stress, activated immune pathways and atherogenicity, ultimately contributing to the depression phenome.

### Insulin resistance, atherogenicity and OMDD features

In this study, we observed that there were no discernible differences in FBG and insulin levels between the NIMETOX cluster and its corresponding control cluster. Prior investigations conducted by the laboratory of the primary author of the present study did not reveal significant alterations in insulin or FBG in OMDD (e.g.,[28]). However, certain studies have indicated a rise in insulin resistance associated with MDD [75]. Nevertheless, it is noteworthy that both FBG and insulin exhibited a negative impact on LCAT and RCT activity, and that insulin or FBG were significantly correlated with various dimensions of the depression phenome.

Insulin resistance is widely recognized as a significant predictor of atherosclerotic disease, as evidenced by the findings of [76, 77]. For instance, within the context of the “insulin resistance syndrome,” it is evident that insulin resistance serves as a causal factor contributing to the emergence of cardiometabolic disturbances [78]. The elevation of insulin levels associated with depressive characteristics may suggest that compensatory hyperinsulinemia could contribute to the features of OMDD, potentially through the activation of inflammatory genes and the promotion of atherogenic processes [78]. Conversely, it is important to note that insulin resistance can also be triggered by oxidative processes that influence insulin signal transduction, the expression of glucose transporter type 4 (GLUT4), and the functionality of pancreatic β-cells [79].

Moreover, the elevation of AOPP production has the potential to induce insulin resistance through the reduction of Akt phosphorylation, which is a critical mechanism in the insulin signaling pathway [80]. Individuals diagnosed with Type 2 Diabetes Mellitus and exhibiting heightened insulin resistance may experience the onset of depression as a direct result of this increased insulin resistance. The implications of insulin in this context can exacerbate atherogenic processes and, in turn, influence other pathways associated with non-insulin-mediated effects on metabolism.

### Interrelated NIMETOX pathways and the features of OMDD

This investigation has demonstrated that there exist highly significant intercorrelations among the various NIMETOX pathways, particularly regarding atherogenicity and LCAT/RCT, juxtaposed with elevated levels of IM-NT, IRS activation, AOPP, insulin, and FBG, alongside notably diminished CMPAase levels. Furthermore, the interplay of these pathways, along with the diminished protective effects of IM-NP and increased ACEs, accounts for a significant portion of the variability observed in the clinical manifestations of OMDD. We have previously discussed that the effects of ACEs on the depression phenome may be partly mediated by activated immune pathways and atherogenicity and RCT biomarkers [14, 81]. In prior papers, we have elucidated that immune activation during the acute phase of MDD exhibits a significant correlation with various facets of the phenome of MDMD and OMDD [10, 32]. In a similar vein, heightened oxidative stress, rather than insulin resistance, was found to be correlated with OMDD [28].

In the present investigation, we observed that the features of OMDD in individuals devoid of MetS exhibited a robust correlation with ACEs and biomarkers indicative of atherogenicity. Conversely, in individuals presenting with MetS, the most significant input variables identified were IM-NT and insulin levels. The interrelationships between lipids and the characteristics of depression present a complex challenge for interpretation, particularly when patients with MetS are included in the analysis [46]. The elevated levels of atherogenicity observed in the MetS group contribute to an increase in variance within the data, thereby resulting in greater imprecision and bias [6].

#### Limitations

As previously articulated, the assessment of additional pertinent NIMETOX pathways has the potential to enhance the accuracy of the established NIMETOX pathway and its correlation with the OMDD features. It is imperative that future research should scrutinize the NIMETOX pathways within the context of other phenotypes of depression, particularly during the acute and remitted phases of MDMD and SDMD.

The principal inquiry pertains to the identification of the primary NIMETOX disorder that gives rise to these interconnected NIMETOX pathways and the associated OMDD features. It is important to note that, in first episode SDMD without MetS, we observed no indications of IRS activation; instead, there was a reduction in CIRS activities alongside an elevation in free cholesterol and ApoE, as well as indirect effects stemming from diminished LCAT activity [20]. The findings indicate that first episode SDMD represents a pre-atherogenic state, even in the absence of MetS or preclinical MetS. Such aberrations may contribute to activated immune and oxidative pathways, resulting in an increase in IM-NT and atherogenicity and perhaps depression and MetS. A lowered prevalence of the PON1 R/R 192 genetic variant, which is accompanied by higher CMPAase activity, and the consequent lower activities of CMPAase in the NIMETOX cluster may further fuel this chain of events. Consequently, one might propose a working hypothesis suggesting that the diminished activity of LCAT and RCT instigates the activation of the other NIMETOX pathways, thereby contributing to the depression phenome.

However, in principle, the cascade of events leading to depression may commence with any pathway that results in deviations within the other pathways. For instance, a significant oxidative stress response may coincide with immune activation, resulting in elevated triglycerides, diminished HDL levels, and increased insulin resistance. This sequence of events may subsequently contribute to the oxidation of HDL and LDL and provoke autoimmune responses, thereby amplifying immune-inflammatory reactions [5].

#### Summary

**Figure 1** encapsulates the findings of the current study. The NIMETOX pathways exert cumulative effects on the features of OMDD. In individuals with MetS, there exist supplementary influences of activated immune networks, as well as insulin levels, on these characteristics. In populations devoid of MetS, there exist pronounced influences of atherogenic pathways on the severity of the associated features. It appears that depressive symptoms in OMDD are caused by intertwined NIMETOX pathways that may exert differential effects depending on whether MetS is present or not. The transition from preclinical MetS to its clinical MetS manifestation may indeed be associated with an increased incidence of depressive features. It is essential to note that a minimum of 78.8% of MDD patients exhibit anomalies in NIMETOX pathways, and this percentage is likely an underestimation, as incorporating other biomarkers could enhance accuracy. Consequently, it is possible that all MDD patients exhibit anomalies in the NIMETOX pathways, which we consider the key disorder in MDD. Furthermore, as previously outlined [11, 46], NIMETOX data and clinical evaluations of the depression phenome should be analyzed using the Research and Diagnostic Algorithmic Rule (RADAR) and the precision nomothetic psychiatry approach to compute quantitative scores for ROI, suicidal behaviors, the patient’s lifetime trajectory, current phenome severity, and pathways. These methods not only improve accuracy but can also be employed to present clinical and biomarker data in RADAR graphs [11, 46]. The latter serve as a fingerprint for patients, reflecting their lifetime and current state, together with the relevant biomarkers that should be used as drug targets to treat the individual patient.

**Figure 1.**
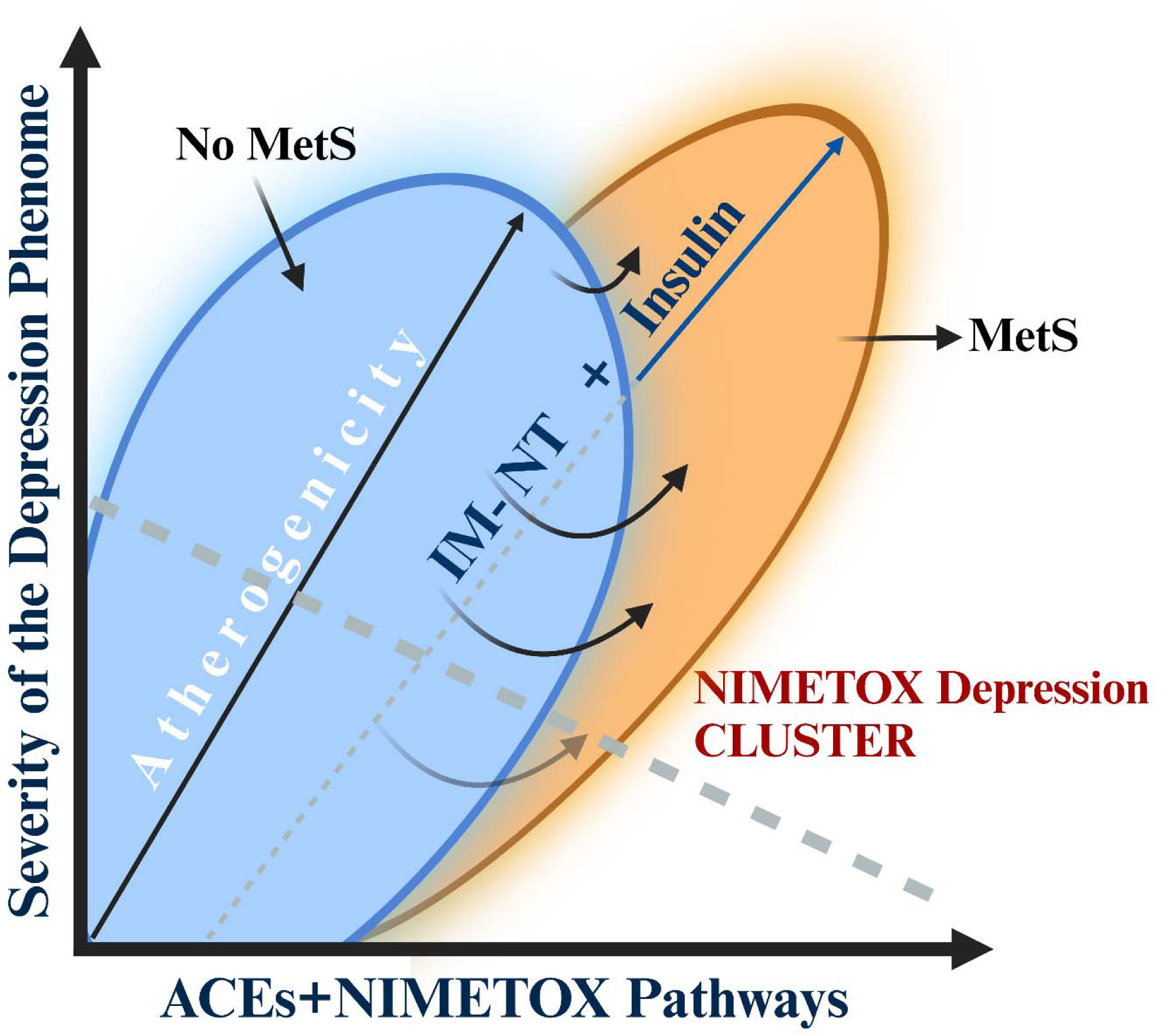
Summary of the findings of the current study. Major depressive disorder in outpatients (OMDD) is characterized by neuro-immune-metabolic-oxidative stress (NIMETOX) pathways and increased adverse childhood experiences (ACEs). Depending on the presence of Metabolic Syndrome (MetS), these NIMETOX pathways may exert additional effects. Thus, in MetS increased immune-linked neurotoxic network and insulin levels may aggravate severity of illness. When no MetS is present, atherogenicity pathways have additional effects on the severity of OMDD.

## Author’s contributions

MM and JK were responsible for the conceptualization and study design. MM also wrote the first draft. MM conducted the statistical analysis. All authors contributed to the editing process. JK was responsible for the recruitment of patients. The final version of the manuscript was authorized by all authors.

## Ethics approval and consent to participate

The Institutional Review Board of Chulalongkorn University’s institutional ethics board approved the research project (445/63). Prior to participating in the investigation, all patients and controls provided written informed consent.

## Data Availability Statement

The dataset produced and/or examined in this study will be accessible from the corresponding author upon reasonable request, following its complete utilization by the authors.

