## supplementary file for "Key factors underpinning neuroimmune-metabolic-oxidative (NIMETOX) major depression in outpatients: paraoxonase 1 activity, reverse cholesterol transport, increased atherogenicity, protein oxidation, and differently expressed cytokine networks"

**ELECTRONIC SUPPLEMENTARY FILE (ESF).**

ESF, table 1. The composite indices that represent the outpatient major depression (OMDD) phenome

| **Index** | **Computation** | **Indicates** |
| --- | --- | --- |
| **DYSNEUANX** | Lifetime Dysthymia (0/1) + z Neuroticism score + lifetime anxiety disorder (0/1) | Lifetime presence of affective and depression-related symptoms |
| **ROI** | z number of depression episodes + z number of lifetime suicidal attempts + lifetime suicidal ideation (0/1) | Recurrence of illness (staging) |
| **Current phenome** | z HAMD + z BDI + z STAI | Integrated severity score of the current depressive episode |
| **Total SB** | z lifetime number of suicidal attempts + lifetime suicidal ideation (0/1) + current suicidal attempts (0/1) + current suicidal ideation (0/1) | Total suicidal behaviors |
| **OSOD** | Z DYSNEUANX + z ROI + z current phenome + current suicidal attempts or suicidal ideation (0/1) | Overall severity of depression, an integrated score that reflects the severity of lifelong affective symptoms |

HAMD: Hamilton Anxiety Rating scale score; BDI: Beck Depression inventory (BDI); STAI: State and Trait Anxiety Inventoty, state version (STAI)

**ESF, Table 2**. Methods used to assay the biomarkers used in the present study.

| **Assays** | **Method** | **Reference** |
| --- | --- | --- |
| Advanced oxidation protein products (AOPP) | AOPP was quantified in mM of equivalent chloramine T using a microplate reader at 340 nm (EnSpire, Perkin Elmer, Waltham, MA, USA). The inter-assay coefficient of variation is less than 10%. | [48, 51] |
| Nitric oxide metabolites (NOx) | Nitrite and nitrate concentrations using a microplate reader (EnSpire®, Perkin Elmer, USA) at 540 nm (Navarro-Gonzálvez et al., 1998), with results expressed in molarity (M). | [51] |
| Paraoxonase 1 (PON1) status | We examined the hydrolysis rate of phenylacetate at low salt concentrations by assessing the activity of arylesterase (AREase) and chloromethyl phenylacetate-ase  (CMPAase). A Perkin Elmer® EnSpire model microplate reader (Waltham, MA, USA) was employed to assess the rate of phenylacetate hydrolysis at a constant temperature of 25 ˚C during a duration of 4 minutes, comprising 16 measurements taken at 15-second intervals. The activity was quantified in units per milliliter (U/mL) using the phenyl acetate molar extinction coefficient of 1.31 mMol/L cm-1. | [22, 23] |
|  | PON1 Q192R genotypes in the current study: CMPA and phenyl acetate were employed to categorize the functional genotypes of the PON1Q192R polymorphism (PON1 192Q/Q, PON1 192Q/R, and PON1 192R/R) (Sigma, PA, USA). The phenylacetate reaction is performed at elevated salt concentrations, which partially impedes the activity of the R allozyme, facilitating a clearer differentiation among the three functional genotypes. We examined the dominant, recessive, additive, and over dominant models of the PON1 genotype. | [22, 23, 49] |
| Total cholesterol (TC), high-density lipoprotein(HDL), triglycerides (TG) | Alinity C (Abbott Laboratories, USA; Otawara-Shi, Tochigi-Ken, Japan). The coefficients of variation for triglycerides, HDL-cholesterol, LDL-cholesterol, and total cholesterol were 4.5%, 2.6%, 2.3%, and 2.3%, respectively. | [46] |
| Apolipoprotein A1 (ApoA1) and ApoB | Immunoturbidimetric assays that involved the Roche Cobas 6000 and c501 modules (Roche, Rotkreuz, Switzerland). Apo A1 and Apo B exhibited intra-assay coefficient of variation of 1.75% . | [14, 20] |
| Free cholesterol (FC) | Was measured using the Free Cholesterol Colorimetric Assay reagent (Elabscience, cat number: E-BC-K004-M). The amounts of free cholesterol. The intra-assay coefficient of variation was 1.9%. | [14, 20, 46] |
| ApoE | Measured using the Human ApoE (Apolipoprotein E) ELISA reagent. The intra-assay CV value was 4.67%. | [14, 20] |
| Fasting blood glucose (FBG) and insulin | FBG levels were measured using the Alinity C analyzer (Abbott Laboratories, USA), manufactured in Tochigi-ken, Japan. The assay employed an enzymatic method utilizing hexokinase and glucose-6-phosphate dehydrogenase (G-6-PDH), with an inter-assay coefficient of variation of 1.7%. Insulin levels were measured using the IMMULITE 2000 system (Siemens Healthcare Diagnostics Products Ltd., United Kingdom, manufactured in Gwynedd, United Kingdom. The assay employed a solid-phase, enzyme-labeled, chemiluminescent immunometric method. | - |

**ESF, Table 3**. Construction of biomarker composites.

| **Index** | **Composition or computation** | **Reference** |
| --- | --- | --- |
| Immune-related neurotoxicity (IM-NT) | First principal component (PC) extracted from sIL-1RA, IL-4, IL-9, IL-17, CCL4, PDGF, CCL5, CXCL12, TNF-α, and TNF-β. The protein-protein interaction (PPI) network based on these cytokines/chemokines/growth factors is enriched in the following GO functions: chemotaxis, defense response, immune response, and response to stress | [32] |
| Immune-related neuroprotection (IM-NP) | First PC extracted from CCL3, CSF1, VEGF, NGF, and IL-10. Its PPI network is enriched in: regulation of neuron death, regulation of microglial cell migration, positive regulation of neurogenesis. | [32] |
| Immune-inflammatory response system (IRS) | Index of the immune-inflammatory response system. Computed as a z unit based composite score, namely sum of z transformations of IL-1α, IL-1β, IL-6, IL-12p70, IL-15, IL-16, IL-17, IL-18, CCL2, CCL3, CCL4, CCL5, CCL7, CCL11, CXCL1, CXCL8, CXCL9, CXCL10, IL-2, IFN-α, IFN-γ, TNF-α, TNF-β, TRAIL, GM-CSF, M-CSF, G-CSF, SCGF | [32] |
| Compensatory immunoregulatory system (CIRS) | Index of the compensatory immunoregulatory system. Computed as a z unit based composite score, namely sum of z transformations of IL-4, IL-10, sIL-1RA, sIL-2R | [9, 12, 47] |
| z IRS – z CIRS | Ratio between IRS versus CIRS | [9, 12, 47] |
| Tumor necrosis factor (TNF) signaling | z Tumor necrosis factor (TNF)-α + z TNF-β + z TNF-related apoptosis-inducing ligand (TRAIL) | [32] |
| z CMPAase – z AREase | Indicates the ratio between the two enzymatic sites of paraoxonase 1, namely CMPAase versus AREase | [49] |
| Insulin resistance (IR) index | z Fasting blood glucose (FBG) – z insulin | [28] |
| Esterified cholesterol ratio (LCAT activity) | Computed using the formula (1-free cholesterol / total cholesterol) x 100 | [7] |
| ApoB / ApoA | Ratio of Apo B on Apo A concentrations | [46] |
| Castelli risk index 1 | Total cholesterol / HDL cholesterol | [5, 6] |
| Atherogenicity index of plasma (AIP) | Triglycerides / HDL cholesterol | [5, 6] |
| Reverse cholesterol transport (RCT) index | z HDL-cholesterol + z LCAT + z ApoA1 + z CMPAase | [5, 6] |
| Comprehensive atherogenicity index (CAI) | z ApoB + z triglycerides + z free cholesterol | [5, 6] |
| z CAI – z RCT | Index of the ratio between CAI and RCT | - |
